# Tuberculosis Mortalities Among In-patients at a Tertiary Hospital in Zambia Between 2018 and 2019 - The Spectrum of Clinical Presentations

**DOI:** 10.1101/2023.01.04.23284196

**Authors:** Webster Chewe, Namakando Liusha, Abidan Chansa, Peter Mwaba

## Abstract

Tuberculosis (TB) has remained one of the most important public health diseases and a leading cause of mortality from a single infectious agent in the world. In-patient mortalities have remained relatively high despite massive investment towards TB elimination. This prompted us to undertake a TB mortality review aimed at understanding the spectrum of clinical presentations in TB mortalities among in-patients in a local hospital set up in Zambia.

Files of 74 in-patient TB related mortalities that had occurred at Kitwe Teaching Hospital over a 12-month period between June 2018 and June 2019 were audited using a structured questionnaire. The descriptive data was analyzed using SPSS v 16.0 statistical software and Microsoft excel 2016.

The audit revealed that 50 (67.6%) of the files were for male patients with a mean age of 39.2 ± 11.6 years. 60(80%) were HIV positive, 60(80%) resided in high-density residential areas. On presentation to hospital, the commonest symptoms included productive cough and fever [31(41.9%) and 30(40.5%) respectively]. HIV positive male patients were presenting in hyperdynamic state (mean pulse rate of 117.2 ± 32.4 per min). Other findings included signs of multi-organ involvement [hypoalbuminemia 9(12.2%), deranged renal function 8(10.8%) and deranged liver enzymes 9(12.2%)] before mortality occurred.

The spectrum of clinical presentations among in-patients with TB in a tertiary hospital include the following; male gender, age younger than 50 years, being HIV positive, residing in a high-density residential area and presenting with unstable hemodynamics. There is a need to focus strategies targeted at strengthening early recognition of clinical instability among admitted TB patients for at-risk populations, including young to middle aged males who are HIV positive.

## BACKGROUND

Tuberculosis (TB) has remained the leading cause of mortality from a single infectious agent in the world despite being a curable and preventable disease^1^. According to the World Health Organization (WHO), 2019 global TB report, an estimated 10 million people fell ill with TB worldwide [5.7 million were males and 3.2 million were females] while a total of 1.5 million people died from TB in the same year^1^. The TB incidence rate has continued to decline at an average of 2% over the recent years and currently stands at 132 (118–146) per 100,000 population^1^. The majority of patients dying from TB were co-infected with human immunodeficiency virus (HIV) while the meta-analysis by de Almeida *et al*. 2018 had shown that in-hospital TB mortality was largely associated with malignancy as the largest confounder^1,2^. The risk of developing TB is estimated to be between 15–21 times higher in people living with HIV than among those without HIV infection^3^.

Zambia is ranked among the top 30 countries with the highest TB burden. In 2018, the total TB incidence rate was 346 (225–493) per 100,000 population^1^, almost 2.5 times the average global TB incidence rate. The HIV-positive TB incidence rate was 205 (133–293) per 100 000 population with the mortality rate among HIV-negative TB populations standing at 28 (16–42) per 100,000 populations and HIV-positive TB population was at 74 (48–107) per 100,000 population^1^. The organization of the healthcare system regarding the fight against TB in Zambia is such that most of the patients that suffer from TB are cared for at primary health facilities that include the health centres, first level clinics and community structures dotted around the districts. However, patients that present with complicated or advanced TB infection are referred to secondary or tertiary level hospitals^4^. It is at these secondary and tertiary level hospitals where the majority of TB related mortalities are recorded.

TB primarily affects the lungs and the severe forms have a tendency to spread to secondary sites in the form of extrapulmonary TB. This disseminated form of TB is difficult to diagnose and increases the likelihood of missing some cases of TB especially at primary care level facilities where the diagnostic capabilities are limited and hence may delay the patients to be referred to the general and tertiary hospitals^5–10^. The WHO defines TB mortality or death as the number of TB patients dying during treatment, irrespective of the cause^11^. This calls for concerted efforts through a multidisciplinary effort to end the preventable causes of TB mortalities as studies have shown that the all-cause of mortality among TB patients were due to aging and having comorbidities like malignancy, liver cirrhosis and renal failure. Whereas presenting with cavitary, miliary and pneumonic radiographic patterns also contributes significantly to TB-related deaths in both developed and developing countries^2,12–14^.

Kitwe Teaching Hospital (KTH), a tertiary hospital that provides health services in the northern part of Zambia has a variety of diagnostics used to diagnose TB like radiographic imaging, sputum acid-fast bacilli (AFB) smear, Xpert MTB/RIF assay and urine lipoarabinomannan (LAM). In a study conducted at KTH by Chanda et al, mortality from TB accounted for 21% of all mortalities and it was the leading cause of mortality among adult patients admitted to the Internal Medicine department^15^. This high mortality prompted us to undertake this audit aimed at understanding the spectrum of clinical presentations in TB mortalities among admitted patients in a local hospital set up, as this will help with early and timely interventions that are both practical and effective in preventing the majority of mortalities caused by TB and improve healthcare delivery to the general populace.

## METHODS

This was an audit of 74 mortality files attributed to TB that had occurred at KTH over a 12-month period between June 2018 and June 2019. Patient files were reviewed using a standardized data collection form. The information obtained included demographic information, clinical symptoms, HIV serostatus, duration of hospital stay and outcome, diagnosis and comorbidities at time of death, radiological and laboratory investigations findings.

The inclusion criteria included all files for patients aged 16 years and older who were admitted and died during the period under review. A total of 30 files were excluded due to the fact that some mortalities occurred within 24 hours of admission and while others had no clear clinical or laboratory confirmation of TB diagnosis (i.e positive genexpert, AFB microscopy, LAM, sputum culture or radiological description consistent with TB diagnosis given by radiologist or experienced physician).

### Data analysis

Patient data that was extracted from the files were recorded and analyzed using Statistical Package for Social Sciences (SPSS) version 16.0 for windows (SPSS Inc, Chicago, IL, USA) while basic descriptive statistics using frequency, percentages, the mean, standard deviation, tables and graphs were analyzed and summarized using Microsoft Excel 2016.

## RESULTS

### Demographic and Clinical variables

A total of 74 patient files with TB related mortalities were analyzed at KTH during the period under review from June 2018 and June 2019 (Table 1). 50(67.6%) of the mortalities occurred among male patients while the mean age was 39.2 ± 11.6 years. Patients from high-density residential areas accounted for 62(83.8%) whereas those from the low density residential areas were 3(4.1%). HIV positive patients were notably high at 60(81.1%) [males 38(51.4%) and females 22(29.7%)] and 4(5.4%) of the patients had unknown or undisclosed HIV sero-status at the time of death.

**Table 1:**
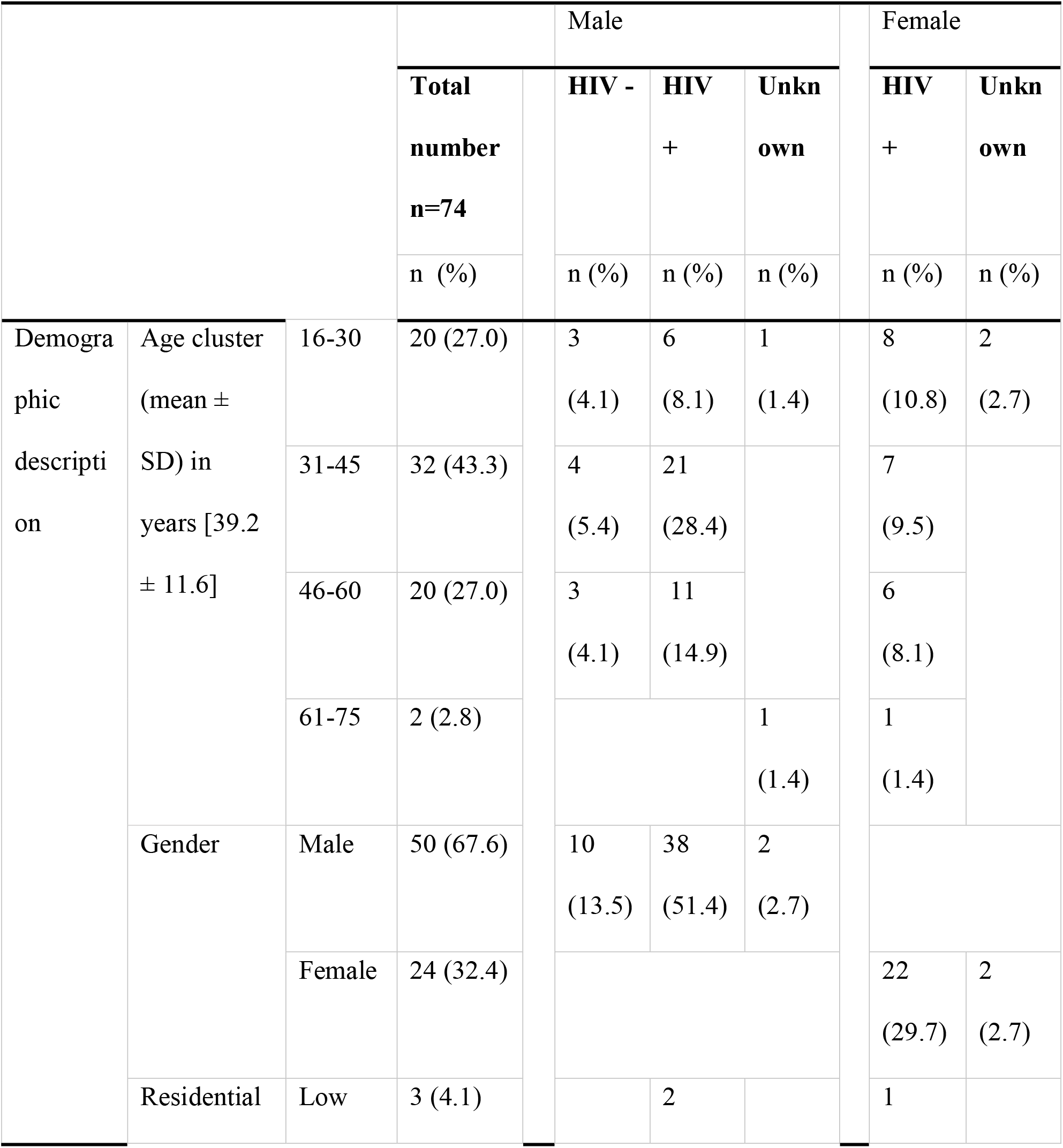
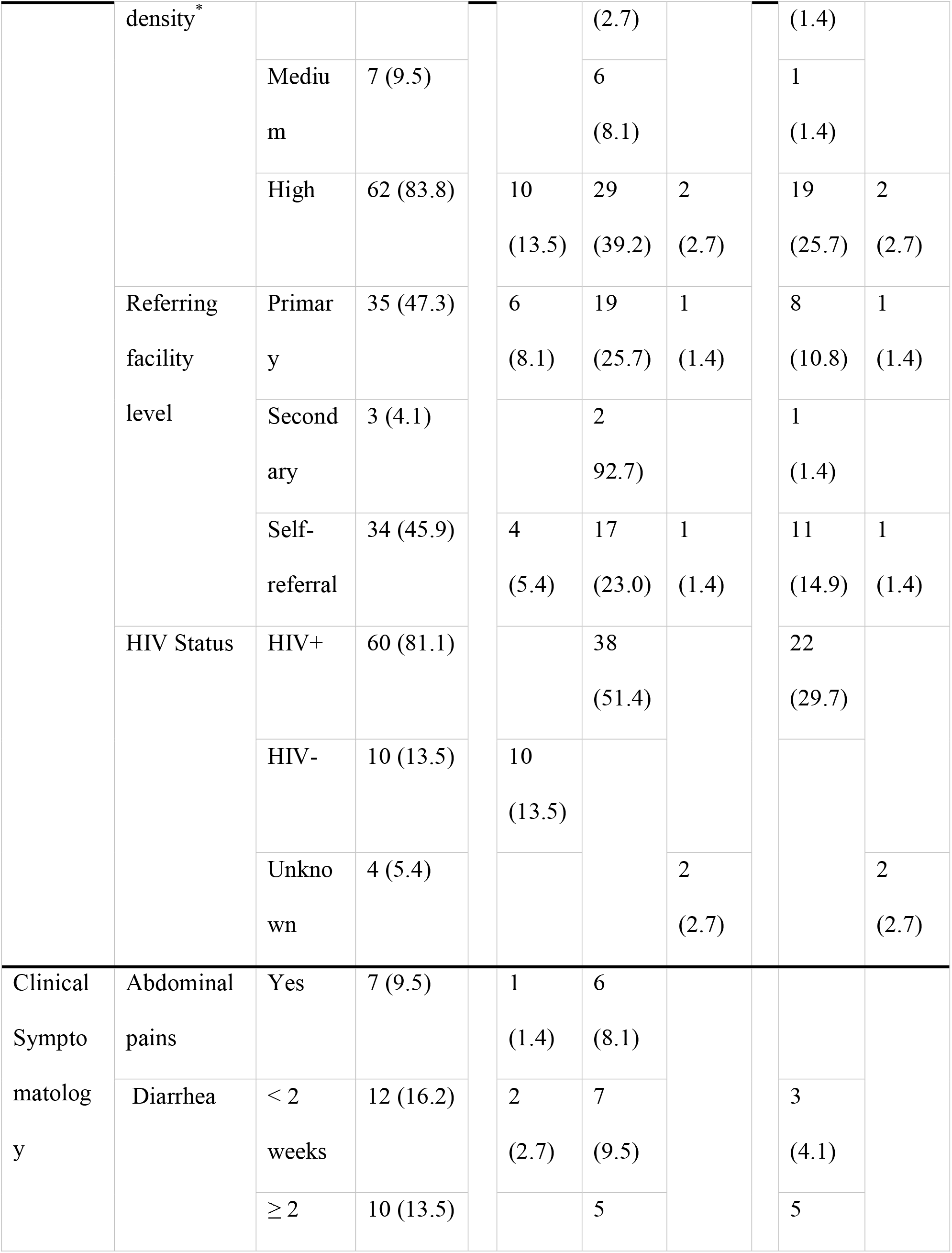
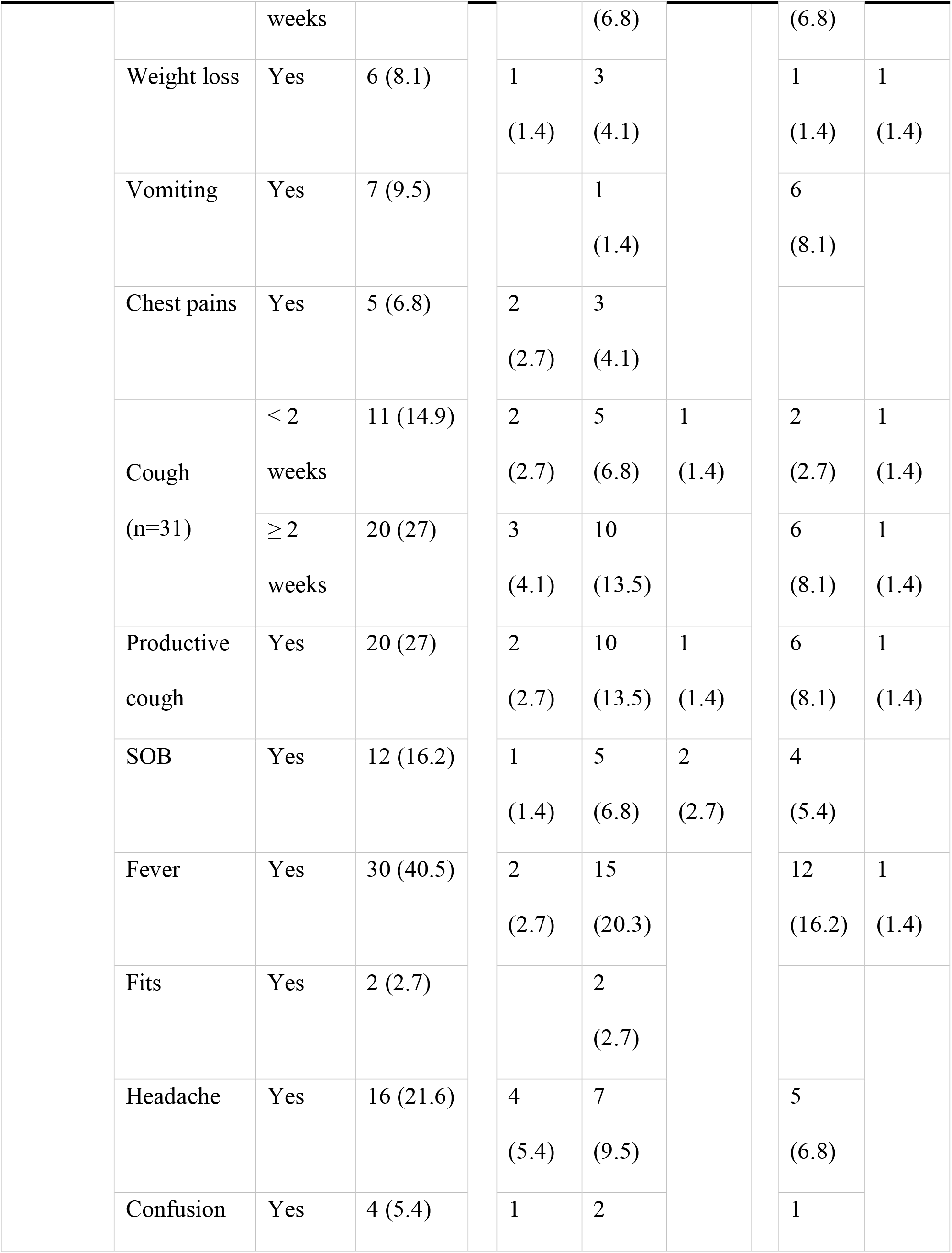
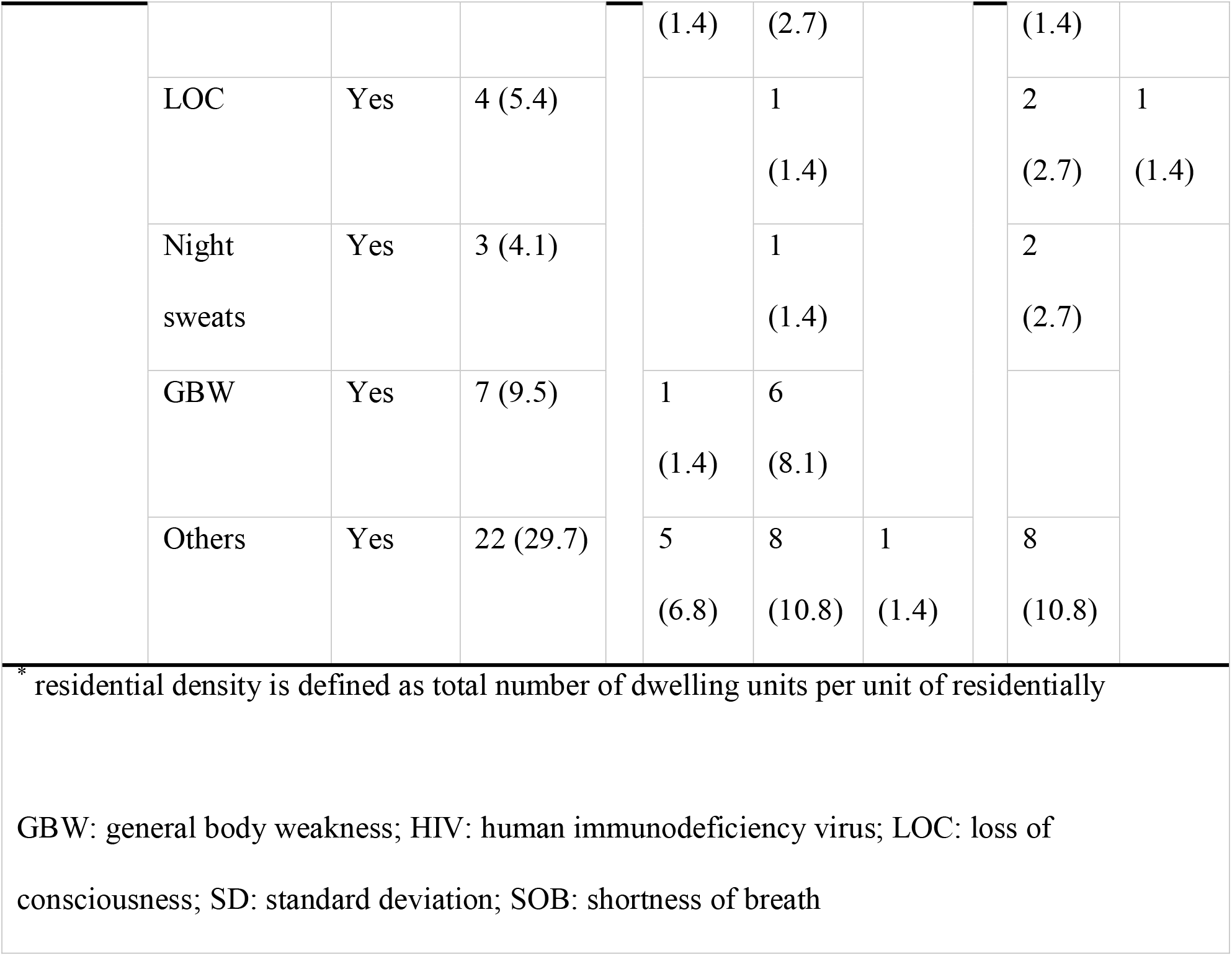
Demographic and clinical characteristics.

The symptoms at presentation included cough 31(41.9%) [11(14.9%) had a cough for less than two weeks while 20(27.0%) had a cough of two-week duration or more]; productive cough 20(27.0%); fever 30(40.5%); diarrhea 22(29.7%); Headache 16(21.6%); and shortness of breath (SOB) 12(16.2%).

### Vital signs

The vital signs on admission were compared to those recorded on the medical ward and the last measured vital sign before death (3 ± 2 hours) as shown in table 2. The mean of “mean arterial pressure” (MAP) on admission and the last measured MAP (MAP ± SD) were 81.5 ± 17.4 mmHg and 75.9 ± 19.1 mmHg respectively. The mean for pulse rate (PR) on admission was high at 112.1 ± 29.7 per minute compared to the last recorded PR before death at 94.1 ± 23.4 per minute. The mean for respiratory rate (RR) was 22.4 ± 5.8 breath per minute on admission and 21.5 ± 4.1 breath per minute before dying.

**Table 2:**
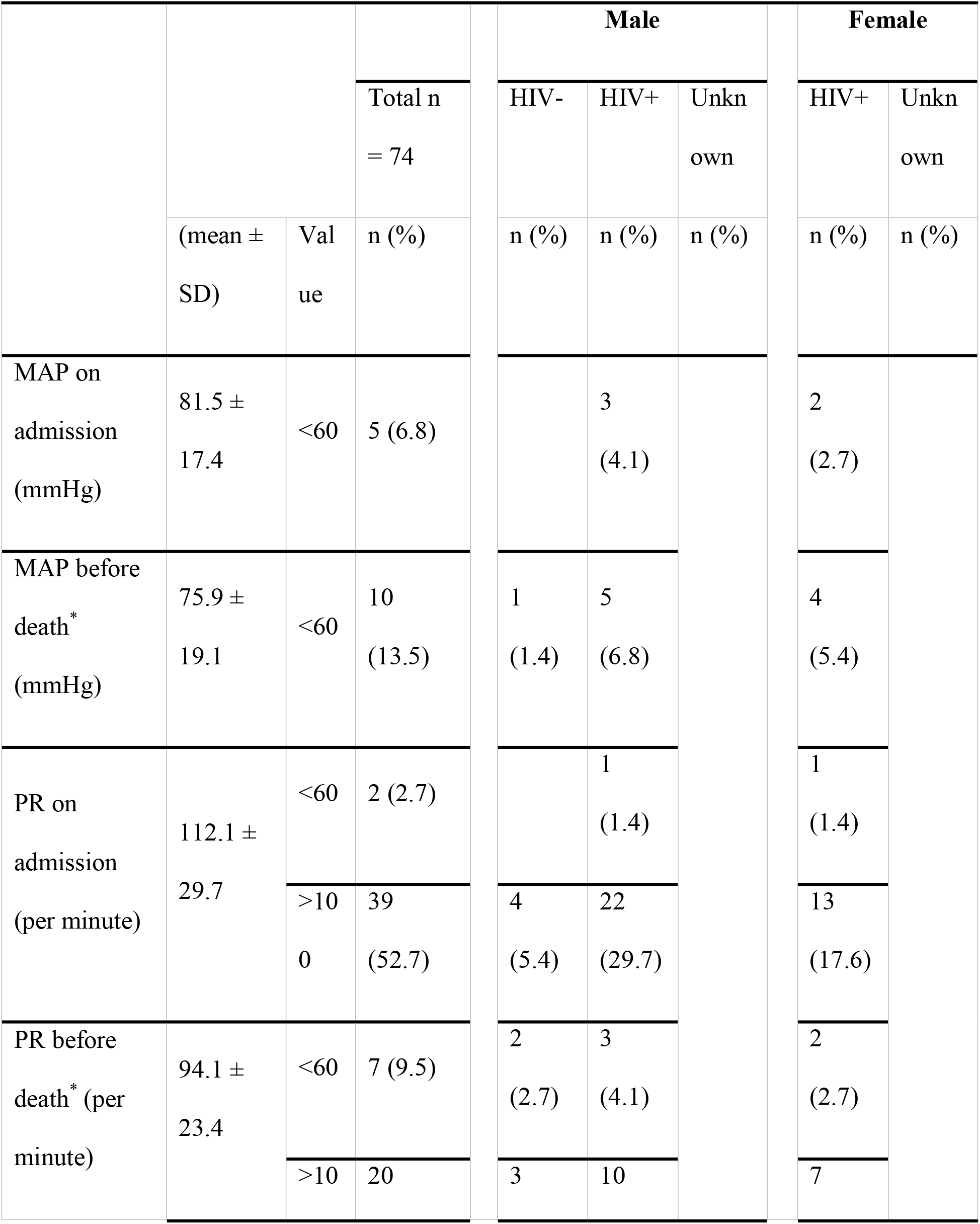
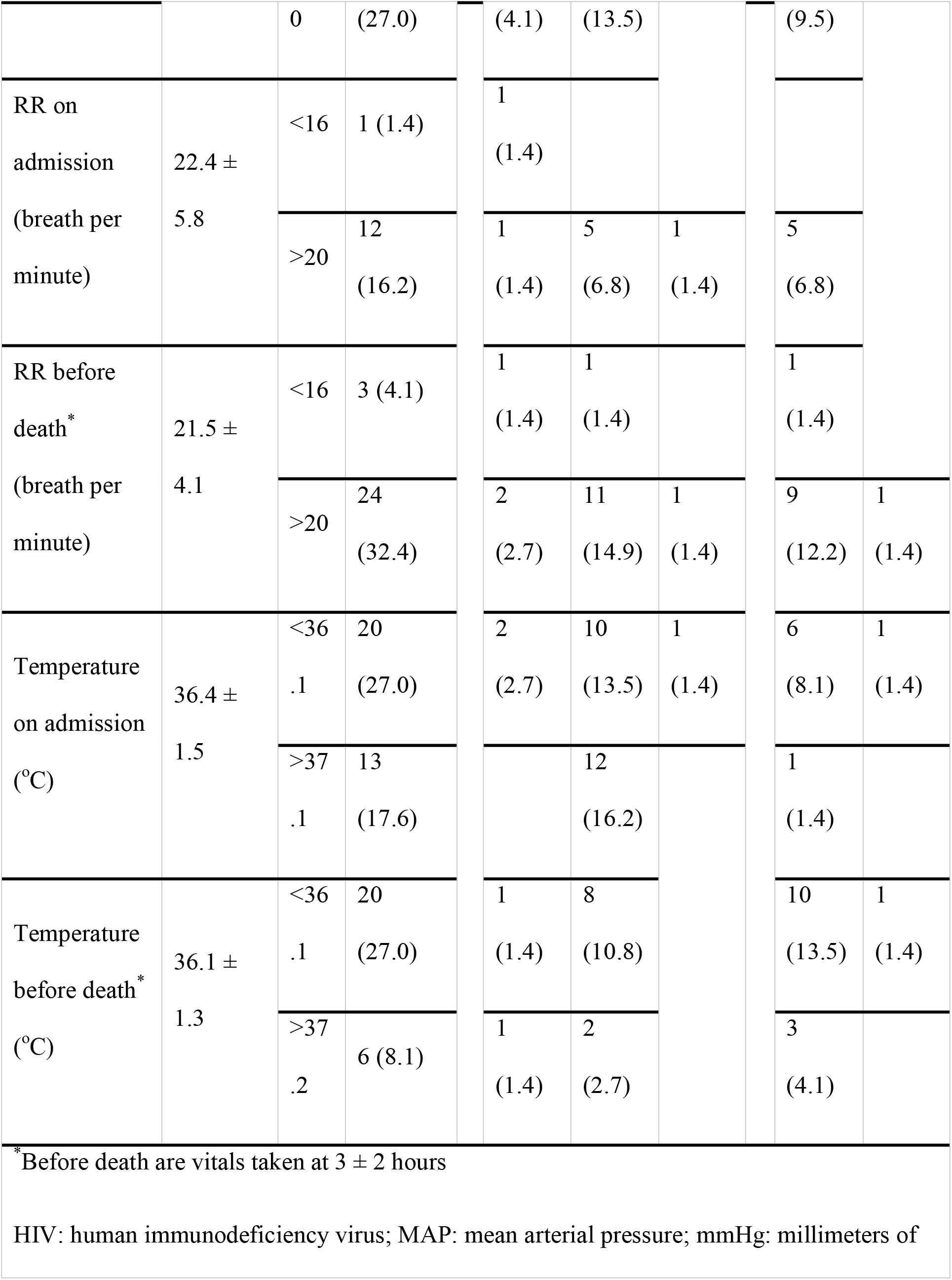
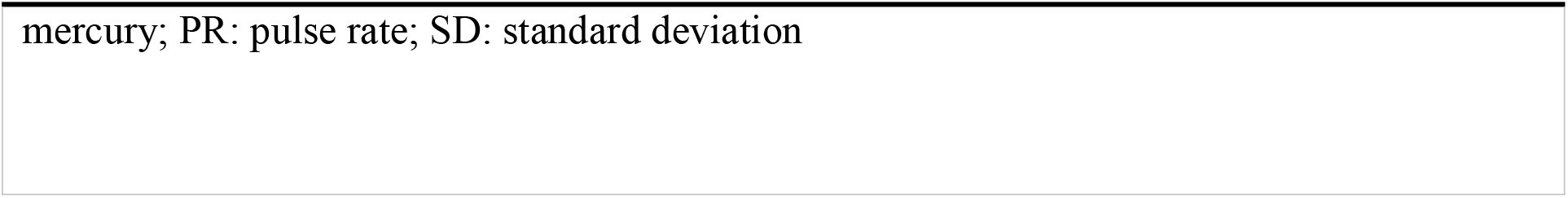
Vital signs

### Laboratory and radiological description

The laboratory parameters (Table 3) showed that 5(6.8%) of the HIV positive males had white cell count (WCC) more than 11.0 × 10^9^/L and 15(20.3%) had hemoglobin (HB) less than 11.5 g/dL. Patients also presented with multi-organ involvement [hypoalbuminemia (albumin less than 35 g/L) 9(12.2%); renal dysfunction (creatinine more than 118 μmol/L) 8(10.8%); deranged liver enzymes (alanine aminotransferase (ALT) and aspartate transaminase (AST) at 9(12.2%) and 14(18.9%) respectively].

**Table 3:**
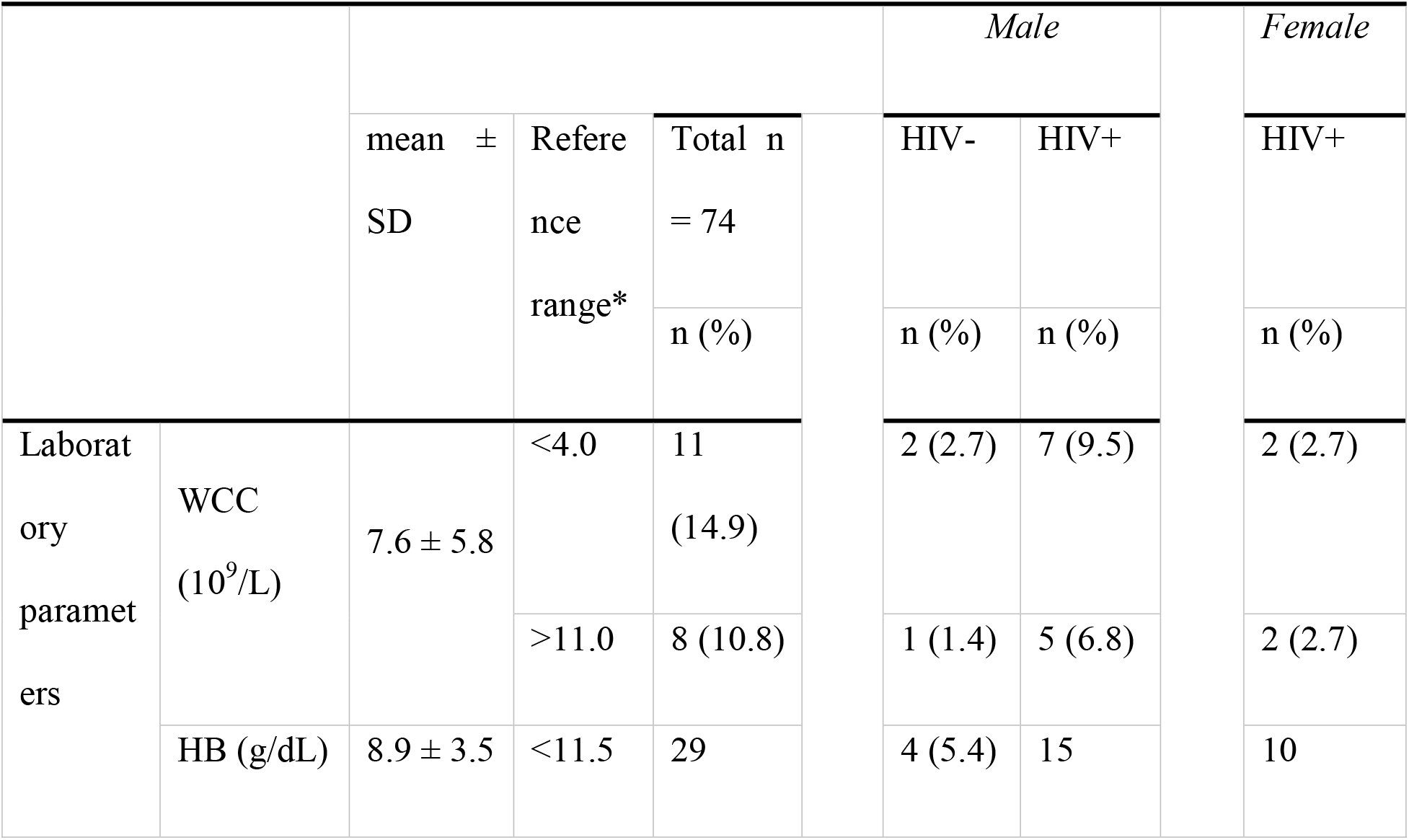
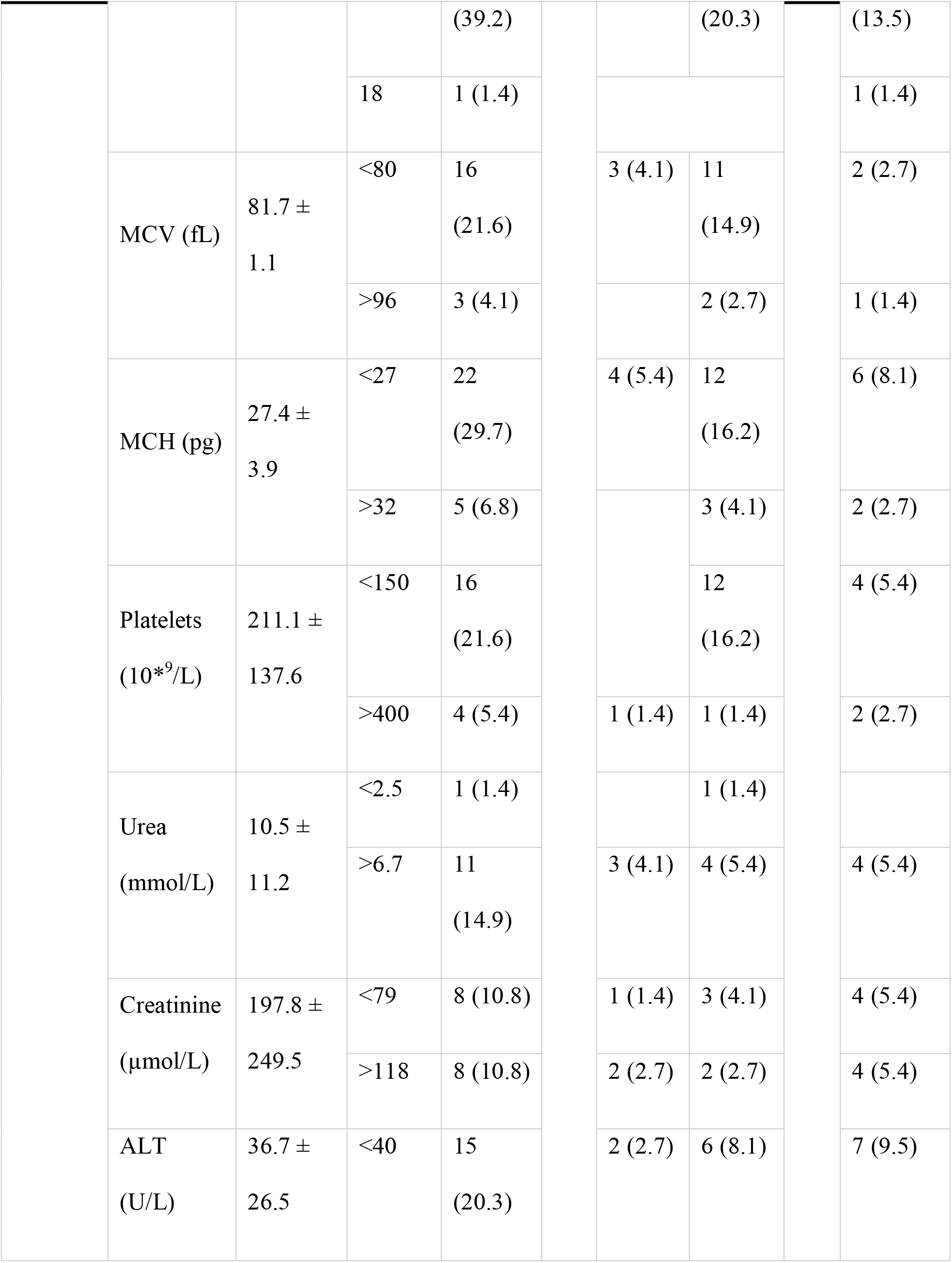
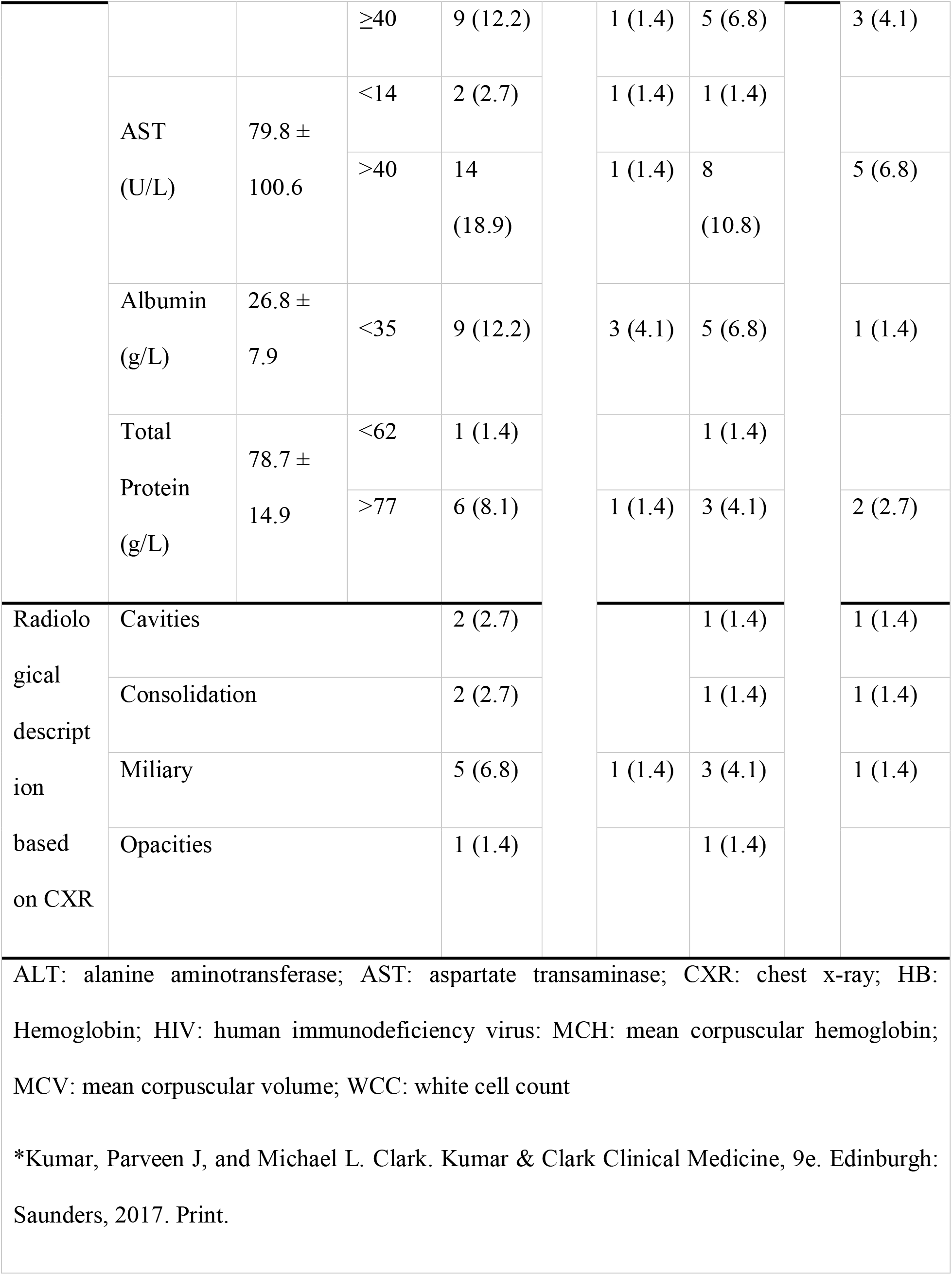
Laboratory parameters and radiological description

## DISCUSSION

In this mortality audit, the majority of the mortalities occurred among males, those younger than 50 years of age and TB/HIV co-infection, consistent with the WHO global TB report of 2019 which showed that TB remains the leading cause of death among people living with HIV^1^. Similar studies have also shown that males who are HIV positive are more likely to die from TB compared to females due to the fact that younger males are difficult to engage during TB treatment and hence have higher default rates^16–19^. These findings reflect the fact that despite resources being channeled towards TB/HIV national programs in terms of active TB case findings and prevention, the follow-up of patients with active disease have not been prioritized and tailored towards the population younger than 50 years of age.

This TB mortality audit further demonstrated a known public health challenge of residing in a high-density residential area and overcrowding as one of the biggest drivers for TB infection. This was consistent with studies that have shown that TB thrives in overcrowded places with most of the high-density residential areas having a high TB/HIV co-infection prevalence^20–22^.

Our audit further showed that the majority of the patients presented with two of the four constitutional TB symptoms. This is consistent with the WHO’s diagnostic screen for TB in people living with HIV^23–26^ which has a negative predictive value of 98%^25^. Diarrhea was also a major presenting feature in this audit. The TB/HIV co-infection makes it difficult to delineate whether the diarrhea is because of TB or HIV as diarrhea has been associated with advanced immunodeficiency. An earlier study by Lucas et. al. had shown no association between chronic diarrhea and tuberculosis nor between diarrhea and tuberculosis of the intestine^27^.

The hyperdynamic states denoted by raised pulse rate in this audit in the majority of the patients is most likely attributed to the compensatory mechanism and multi-organ involvement^28-31^. A study by Seccareccia *et al* showed that the heart rate was an independent predictor of overall mortality in the middle aged male population^32^ with no specification to patients with TB.

In this TB mortality audit, 39.2% had anemia (HB less than 11.5 g/dL) consistent with studies that have shown that anemia is a common hematologic manifestation in TB patients and a strong risk factor for mortality^33–35^. The low hemoglobin manifestation is not surprising as TB infection being a chronic disease is characterized by release of inflammatory mediators which impair production of erythropoietin, suppress bone marrow response to erythropoietin and alter iron metabolism leading to impaired erythropoiesis^36^. Other hematological manifestations included microcytosis at 21.6% and macrocytosis at 4.1% denoting iron and vitamin B12/folate deficiency respectively.

Our audit also showed multi-organ involvement as evidenced by reduced albumin levels, deranged renal function and liver enzymes. Multi-organ involvement is common in disseminated forms of TB^37–39^. In the metaanalysis by de Almeida *et al*, they found that malignancy was a significant attributor of mortality^2^, this was not the case with our study as we did not look for malignancy.

The limitations of this study was the fact that it was a retrospective TB mortality file review and hence relied on existing data. The validity and reliability depended on the documented information and files extracted from the registry. As a developing nation with electronic file storage still in infancy, some files were either missing or had missing data which may have reduced the supposed number of files reviewed hence may lead to some parameters being either under- or over-estimated. Despite all the above limitations, the strength of the study was that it was conducted on the only tertiary hospital in the second populous city in Zambia and a further clinical study is warranted to understanding the predictors of mortalities among inpatients with TB.

## CONCLUSION

The spectrum of clinical presentations among in-patients with TB in a tertiary hospital include the following; male gender, younger than 50 years of age, being HIV positive, residing in a high-density residential area and presenting with unstable hemodynamics. There is a need to focus strategies targeted at strengthening early recognition of clinical instability among admitted TB patients for at-risk populations, including young to middle aged males who are HIV positive.

## Data Availability

All data produced in the present work are contained in the manuscript

## RECOMMENDATION

We recommend that a follow-up prospective study, in which patients that die from tuberculosis, be conducted to ascertain the predictors of mortality and whether malignancies are a contributing risk factor to the high mortality among admitted TB/HIV co-infected individuals in this setting.

## ABBREVIATIONS

AFB: acid fast bacilli
ALT: alanine aminotransferase
AST: aspartate transaminase
CXR: chest x-ray
GBW: General body weakness
HB: hemoglobin
HIV: human immune-deficiency virus
KTH: Kitwe teaching hospital
LAM: lipoarabinomannan
LOC: Loss of consciousness
LPA: line probe assay
MCV: mean corpuscular hemoglobin
MAP: mean arterial pressure
MCV: mean corpuscular volume
MGIT: mycobacteria growth indicator tube
mmHg: millimeters of mercury
NHRA: National Health Research Authority
PR: pulse rate
RR: respiratory rate
SD: standard deviation
SOB: shortness of breath
TB: tuberculosis
TDRC-ERC: Tropical Diseases Research Centre Ethics Review Committee
WCC: white cell count
WGS: whole genome sequencing
WHO: World Health Organization

## ACKNOWLEDGEMENTS

We wish to extend our profound gratitude to the staff in the department of Internal medicine and management at Kitwe teaching hospital for the permission to gain access to the medical files and subsequent data extraction.

## AUTHORS’ CONTRIBUTIONS

All authors contributed equally to the development of this manuscript.

## ETHICAL CONSIDERATIONS

The ethical clearance was obtained from Tropical Diseases Research Centre Ethics Review Committee (TDRC-ERC), I.R.B. No. 00002911, F.W.A. No. 00003729 under reference number: TRC/C4/12/2020 and National Health Research Authority (NHRA) under reference number: NHRA00007/08/01/202 and NHRA0001/26/04/2021. Further, permission was obtained from management of KTH to conduct this study. Patients’ mortality files were entirely confidential and private information like name and address were protected.

